# How do families cope with major societal stressors: a qualitative study of family coping during the pandemic?

**DOI:** 10.1101/2025.05.08.25327227

**Authors:** Paraskevi Bali, Papoula Petri-Romao, Fiona Turner, Rebecca Dyas, Oliver English, Jack MacLean, Irene O’Neil, Helen Minnis

## Abstract

**Background:** Major societal stressors such as wars, natural disasters and pandemics severely disrupt family life. However, not all families are impacted equally. During the COVID-19 pandemic, surveys highlighted how most of UK society coped, but tended to exclude high-risk families. We sought to redress this.

**Methods:** Forty-three participants (25 parents from high-risk families; 18 family-support professionals) were interviewed about family their experiences during the first three months of lockdown. Interviews were conducted in two phases: around the start of the pandemic in April 2020, and during the first easing of restrictions in July 2020. Interviews were analysed using Thematic Analysis.

**Results:** Six major themes were identified: health & wellbeing, family dynamics, work & employment, education, home environment and adherence to government restrictions. Families faced challenges in creating a balance between parent’s work and childcare. A wide range of risk and protective factors, and the influence of pre-existing experiences, contributed to whether a family entered a ‘Virtuous’ (supportive) or a ‘Vicious’ (maladaptive) cycle of family coping. Negative pre-existing experiences worsened some families’ adaptation but helped other families to have resilience.

**Conclusion:** This study extends the Family Stress Model by illustrating the potential for resilience among high-risk families, highlighting potential mechanisms that might enable some families to transform adversity into strength. These findings may be useful for professionals supporting high-risk families coping with societal stressors and for the development of recommendations for future pandemic preparedness.

## Introduction

Family stress has long been recognized as a significant risk factor for child development challenges and adverse experiences, such as child maltreatment (Garner et al., 2024). Family stress may impact child well-being, and increased stress can disrupt parenting behaviours and attachment security (Wu and Xu,2020), thus influencing developmental outcomes. In particular, acute family stressors, such as covid-19, is linked to poorer mental health outcomes in both parents and children, creating a cyclical effect that further exacerbates the stress and developmental challenges and negative outcomes, within these households such as including disruptions in parental mental health, reduced emotional availability, and economic strain (Garner et al., 2024) These stressors collectively contribute to adverse developmental trajectories in children (Marthur et al. 2023; Maserik and Conger (2019). Mechanisms through which stress adversely impacts family dynamics and child development are still unclear – we do not yet fully understand why some high-risk families experience a greater impact than other high-risk families, when experiencing the same stressful events.

Previous research has shown that longitudinal designs are essential for capturing the evolving impact of stress over time (Conger et al., 2004). It is challenging to conduct longitudinal research to examine the impact over time of universal, severe, stressors affecting all families, such as situations of war or natural disasters. The COVID-19 pandemic therefore presented a rare occasion to better understand the effects of a prolonged, pervasive, global stressor on family functioning and child development. Unlike typical stressors, the pandemic imposed widespread economic and psychological strain across diverse populations, providing a unique context to explore stress mechanisms and their developmental implications longitudinally. Measures intended to reduce the transmission of COVID, including ever-changing conditions such as lockdown, social distancing, self-isolation, working from home, and closing of schools, disrupted daily routines and significantly impacted family wellbeing (Bayham, 2020). Reports have equated the psychological impact of the pandemic to that of natural disasters and the World War II aerial bombing campaigns in the UK, illustrating its severe and far-reaching effects on population mental health (Pain et al., 2020; Jones, 2020; Fiorelli, 2020; Makwana, 2019; Betancourt et al., 2018).

Recent meta-analyses revealed that one in five children globally are experiencing clinically elevated depression symptoms and one in four children are experiencing clinically elevated anxiety symptoms. It is crucial to understand the effects of serious stressful events and the mitigation strategies for children and parents (Serrano-Allacron et al., 2022; Yamamoto et al., 2022; Joensen et al., 2022 Clayton, 2020; Dodd, 2020). Given the risks that a future pandemic-even from a different virus-may pose for parents’ and children’s mental and physical health, it is important to detect specific factors that may place parents and children at elevated risk. In the last 5 years, several policies have been published suggesting actions to prepare for a future disaster or cross-societal stressful event such as a pandemic https://www.ready.gov/pandemic. Although research has been conducted highlighting the vulnerabilities and needs of specific groups of people (ref), most of the guidelines and research outcomes focus on how to teach people new skills to limit the negative impact of the disaster on society and people’s health. Here, we analyse the experience of “high-risk families” with historical disadvantages at two timepoints during the COVID 19 pandemic (Liu et al., 2021)

High risk families are often the most vulnerable to stress and yet are the most difficult to engage in studies, creating gaps in understanding the pandemic’s full impact on the populations most at risk. To address these research gaps, we purposively sampled marginalized families and those experiencing additional stressors pre-pandemic, as well as professionals working with these families. This approach allowed us to investigate, in a longitudinal framework, the compounded effects of the pandemic as a major stressful event and to create a model of adaptation for families in possible future stressful scenarios.

## Methodology

### Participants

We conducted 43 semi-structured, telephone or video interviews with parents, and professionals who work with families, located in Scotland and England in April 2020, shortly after the first lockdown started in the UK. The second phase was a follow-up of 41 interviewees (1 professional and 1 family dropout) from June to July 2020, just after the UK government had eased the lockdown measures. High-risk families with broad-ranging backgrounds were recruited (see Table 1), along with professionals who work closely with vulnerable families, allowing us to involve families not usually represented in surveys and to examine possible contributing factors that are already known to be associated with increased stress/challenge (e.g., history of a mental health condition, living in a foster home).

**Table 1.**
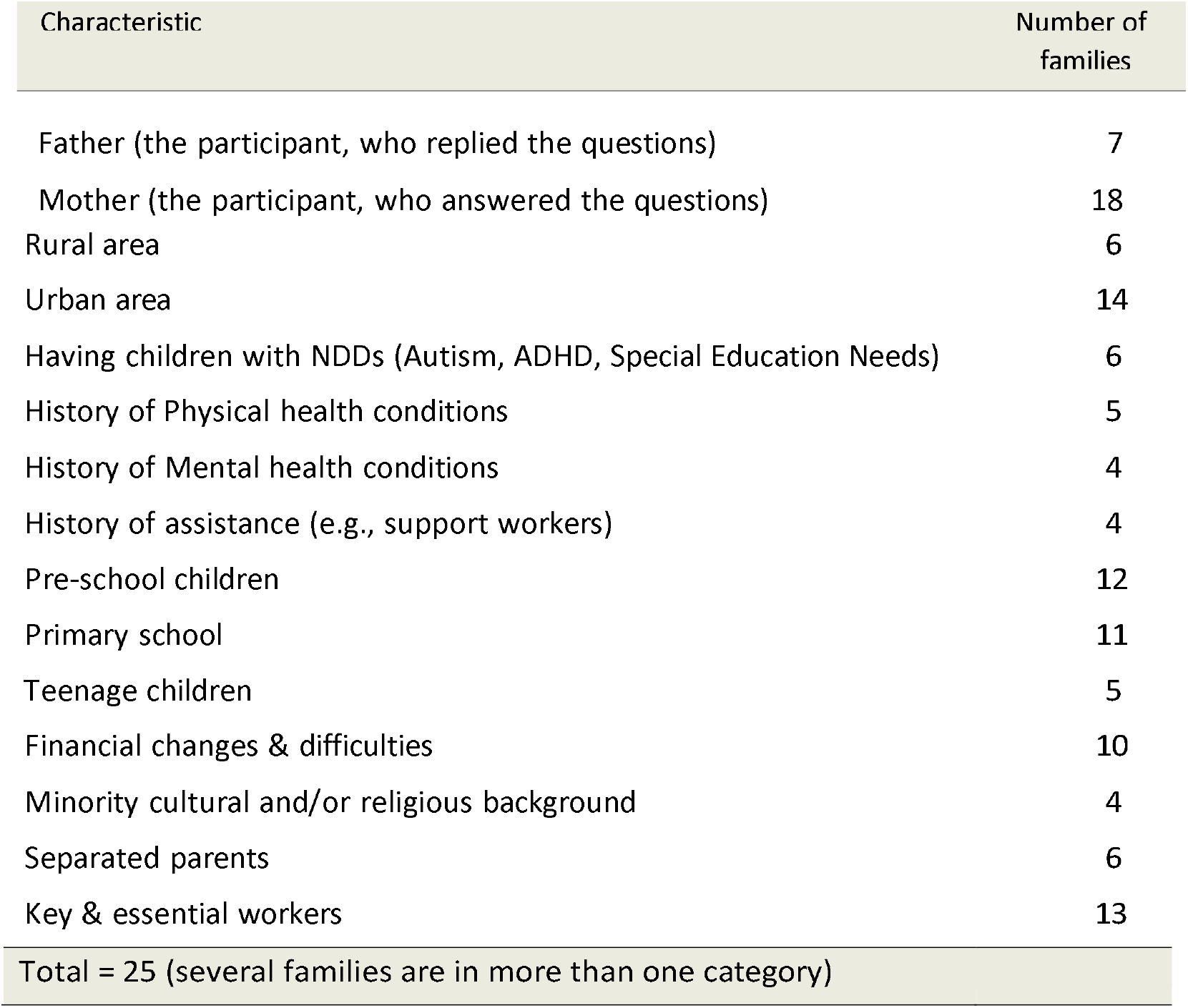
Participants-Families characteristics.

Families:Through our network of health, education, social care, and voluntary sector colleagues, we purposively recruited 25 parents to tell us about their personal experiences of lockdown. We aimed to explore a wide range of different experiences that may have been impacted by the lockdown, so we recruited families with a variety of characteristics, see Table 1.

Professionals:We recruited 18 professionals from Scotland and England: 10 social workers, 3 teachers, 3 nurses, 2 police officers, to give us their experiences of how high-risk families they work with were coping throughout the lockdown and as restrictions began to ease.

### Procedure

Targeted snowball sampling was employed to reach specific populations of interest (e.g., history of a mental health condition, living in a foster home) (Parker, Scott and Geddes, 2019). Semi-structured one-to-one interviews (Jasmhed, 2014) were conducted by four researchers. A topic guide was used to guide the interview process and steer the dialogue, whilst allowing participants to raise their own topics. The questions were designed to be flexible, open-ended and broad. The interviews were conducted online and lasted approximately 1 hour in the first phase, and 40 minutes in the second phase.

### Data analysis

The interviews were audio recorded and transcribed verbatim. Transcripts were then analysed using the guidelines outlined in Thematic Analysis (Braun & Clarke, 2023). Three analysts independently analysed the transcripts of the interviews and extracted themes according to the following process:

The transcripts were checked against audio recordings for accuracy. The three analysts independently created the initial codes and grouped these after agreeing upon a set list of codes. After reviewing the resulting codes, 21 subthemes were identified. The analysis team then grouped these sub-themes into the 6 main themes. We used judging criteria (homogeneity and heterogeneity) to group all data relevant to each potential theme. A further review was conducted of the themes to be sure that the final themes accurately represented the content of the transcriptions. Then, researchers generated thematic maps of the analysis (separate for each phase of interview) and clear definitions and names for each theme. We repeated the same process for both Phase 1 (first interviews) and Phase 2 (follow-up interviews). Finally, an overview of the themes from both phases was taken to generate our final interpretation.

### Ethical considerations

The study was given ethical approval by the University of Glasgow College of Medicine, Dental, Veterinary and Life Sciences Ethics Committee and followed GDPR guidelines.

## Results

### 1. ain themes

Thematic Analysis resulted in 6 main themes: health & wellbeing, family dynamics, work & employment, education, home environment, and adherence to government restrictions/measures, and 21 sub-themes (see Figure 1). A further analysis of the content of the interviews allowed us: a) to do a comparison between the two phases of interviews, b) to develop a model about Virtuous & Vicious cycles and c) highlight the impact of pre-existing factors on it.

**Figure 1.**
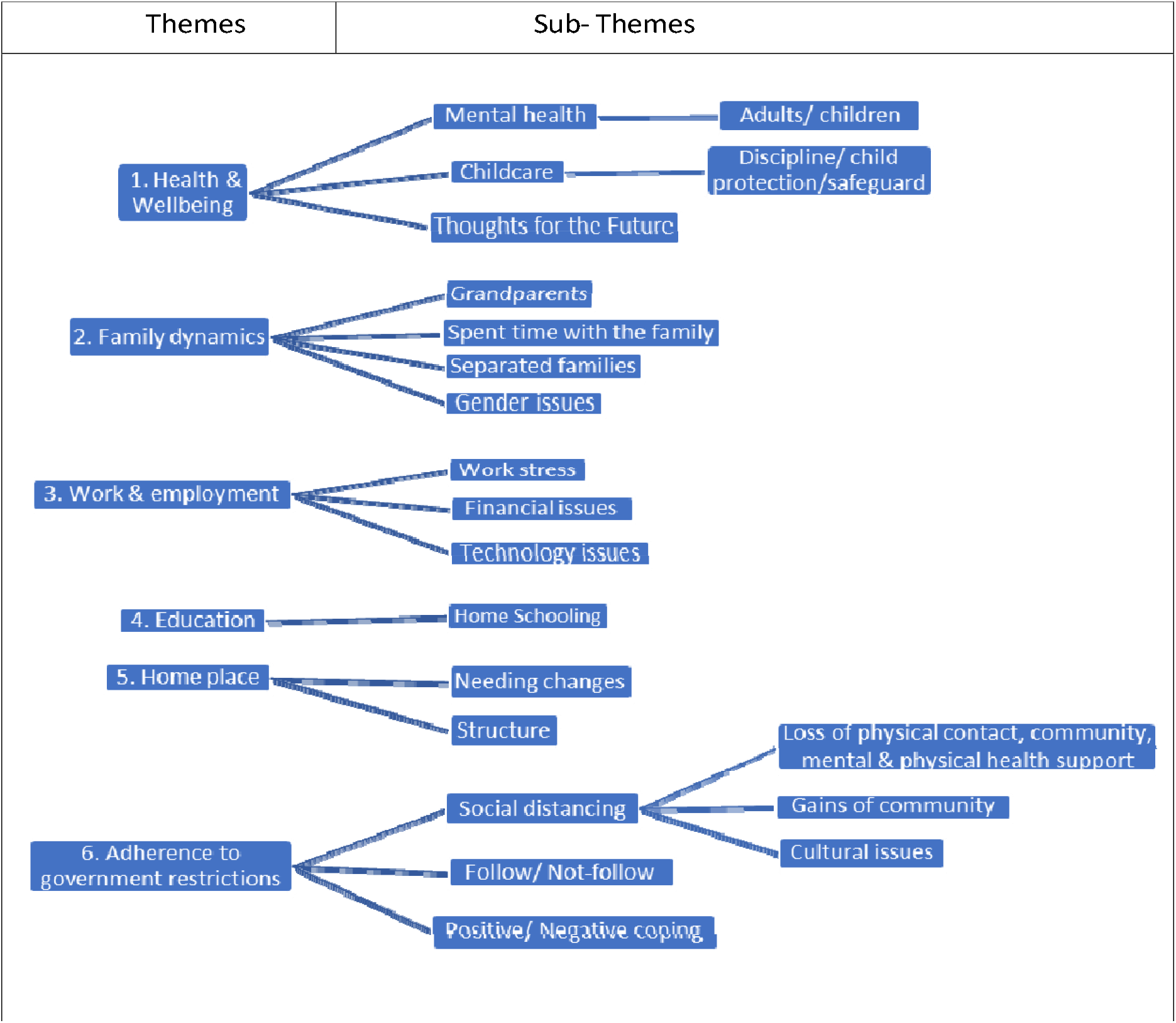
Themes and sub-themes.

#### 1.1 Health & Wellbeing

Parents’ and children’s health and wellbeing fluctuated during the lockdown and easing the restrictions phases. Positive impact: For many families lockdown had a positive impact on their health and wellbeing. Families able to balance work, school, and home-life, and maintain their social interactions, seemed to be more resilient and to have lower stress levels. Families living in rural areas felt less restricted, compared to families in urban areas, who usually felt overwhelmed by the lack of outdoor activities. After easing the restrictions, many families reported better mental health since they had created a functional daily routine and/or they were able to go back to work and meet others face-to-face.

Right now, it is actually really nice to be able to take the time to just go for a walk and hang out and just do some painting, you know just like going into the garden and not worry about what time it is, that is really lovely, so we are enjoying that aspect. (ID16d, family).

Negative impact: of lockdown: Many families suffered from anxiety and worries about the future. Parents/carers felt stress trying to balance work, home-life, and childcare. Families with a history of need for assistance and problematic relationships were more likely to report negative emotions, more conflicts, high anxiety, and difficulties adapting to the changes and follow government restrictions. Children with a history of physical or mental health conditions such as neurodevelopmental conditions (NDC)s were struggling to cope with the changes: they felt socially isolated from their friends, they had sleep problems, emotional, behavioural difficulties and difficulties separating from parents. After easing the restrictions there were parents and children who were exhausted and stressed, experiencing problems with sleep and concentration, feeling unmotivated and with limited ability to engage their coping skills.

Some of our families are struggling because of the nature of the child that they are looking after[..]for children with behavioural difficulties, we are getting a lot of phone calls from families who are absolutely struggling trying to manage their child’s behaviour (Professional -community nurse).

#### 1.2 Family dynamics

The structure of families during the lockdown changed and was often reduced to the core family, without additional support. Lockdown was an opportunity for some families to spend quality time doing activities and communicating with each other. However, some parents and carers had difficulties in applying the discipline methods they used pre-pandemic, creating new rules and reward systems or changing the existing ones to manage children’s behaviour, whereas other parents applied stricter discipline methods. Social workers reported increased domestic violence referrals and children with additional needs were at a higher risk. Foster families reported difficulties to adapt to guidelines and more stress mostly because of the lack of support from services or other sources. Also, some foster-care families with many children experienced overcrowding and associated conflict.

It is nice to all wake up together (…) so mornings are a bit more relaxed you could say (family).

I am using all the strategies and trying de-escalate the situation …I really don’t know how else to do it to be honest, I am at a bit of a loss of strategies for that, the alternative is I let him get his own way…. sorry (family)

…domestic abuse referrals via the police, have gone up significantly. So, in the last month, so for April that was the highest that we’ve had all year[..] but the 19^th^ May we had more domestic referrals than we had for the whole of last month, and interestingly they have all gone up to medium risk referrals rather than the standard risk ones (professional)

#### 1.3. Work & employment

Government guidelines promoting homeworking caused many challenges. Several families faced financial difficulties. Most of the parents had to change their working hours and rearrange/re-organise their homes to be more functional for their work. The increased use of technology for work from home revealed the gap between families, between those who were experienced with technology and able to adapt easily to remote working and those without the appropriate equipment or technological knowledge, who were more anxious and stressed about working remotely. Key workers, who didn’t work remotely, were stressed and worried that they could transmit the virus to their family.

Families who are in poverty, they still don’t have internet at home, they don’t have such resources (, social worker).

I work particularly at night when she (her daughter) is in her bed I am trying to catch up with a lot of things, and possibly doing a lot more hours than I was meant to be doing (parent/professional, social worker).

#### 1.4 Education

Remote schooling caused many changes in families’ daily lives. The majority of the families were struggling with home-schooling. Children without the appropriate IT equipment in their homes and those with neurodevelopmental problems and/or special educational needs were struggling more to adapt to home schooling. However, some families reported that the communication with the teachers was increased because teachers tried to guide to help children with their homework remotely.

I feel wee bit bombarded from both of the schools[..] we feel that the workload that both of the children have had has been an awful lot (social worker)

He (teenage son with autism) doesn’t have the structure, so he is upset and he is frustrated and he is struggling to understand, […], so we are trying to maintain every side of it and that’s just kind of not possible (family with with autistic son).

#### 1.5 Home environment

The layout of homes and the size of the house had an immediate impact on the family’s wellbeing and adjustment to the restrictions. Families with bigger houses with extra rooms and more space felt fortunate, because parents and children could concentrate on work and schoolwork and had more options for activities without feeling crowded. Also, families living near a park and able to visit frequently had a good adjustment during lockdown, reported better mental health, and more balance in their daily tasks. On the contrary, families, who lived in small flats/houses or without access to an open space, were struggling more:

We have got a garden and a big enough house that we have…you know my husband is working in a completely separate place to me (family).

There is a lot of strain on us, because we are in a three-bedroom house, and the fact that we have four kids in the house, and we try and find places that are safe for the kids to play and there is not a lot of places for them (family).

#### 1.6 Adherence to government restrictions

There were families able to follow the government restrictions (e.g., maintaining physical distance, wearing masks and self-isolating), and those who found it challenging. Many families didn’t exactly understand the government’s guidelines and what they had to do, especially during the first month of the lockdown. Families in which parents and/or children had learning difficulties, mental health problems or language barriers had additional difficulties adhering to the restrictions. Social distancing was reported by families as lack of physical contact with their friends and relatives; and lack of community support, because of a reduction in the opportunities for socializing and assistance. However, in some instances there were gains in communities.

After the measures were eased in June/July, there was a new category of people who ignored the easing of restrictions, because they didn’t feel safe going out and meeting others, even within the recommended social distance.

She [her daughter] will have tantrums just over not being able to go out to the shops or go out to nursery and she will take a tantrum because she can’t go near people, that’s the main one that really gets her because she doesn’t understand (single mum family).

I think stress levels and child coping have probably eased actually, yeah, I think they have got better, just as we have adjusted to it and settled into this routine. (family)

### 2. Comparison of 1^st^ and 2^nd^ phases

There was convergence amongst all participants regarding the characteristics of families’ problems and challenges during the lockdown. During the first phase (April 2020), families had to create a balance between work, the requirements of the new technological applications, childcare, adults’ and children’s mental health issues and the consequences of following or not following the lockdown restrictions. Some families were able to adapt, create and keep a new routine more easily than other families, who were facing difficulties in adapting to lockdown restrictions.

During the follow-up and after the easing of the second lockdown (June-July 2020), families tried to maintain the balance they created in previous months or were still struggling to create that balance. Same families worked and built towards better adaptation and to control stressful factors compared to the first weeks of lockdown. At the same time there were those who had adapted to the restrictions, but were very stressed because they were afraid to return to their pre-covid routine. Specifically, families with children with NDCs were struggling more than other families to adjust to the changes and create a new routine in the beginning of lockdown as well as in during the follow-up, after the easing of the restrictions.

### 3. The model of virtuous & vicious cycles of family adaptation to lockdown restrictions

There were, typically, two different trajectories of family coping over time period from initial lockdown to just after the lockdown ended (approximately 3 months). We have described this using a model of “virtuous and vicious cycles” (Figure 2) regarding the adaptation process of families to government restrictions over time.

**Figure 2.**
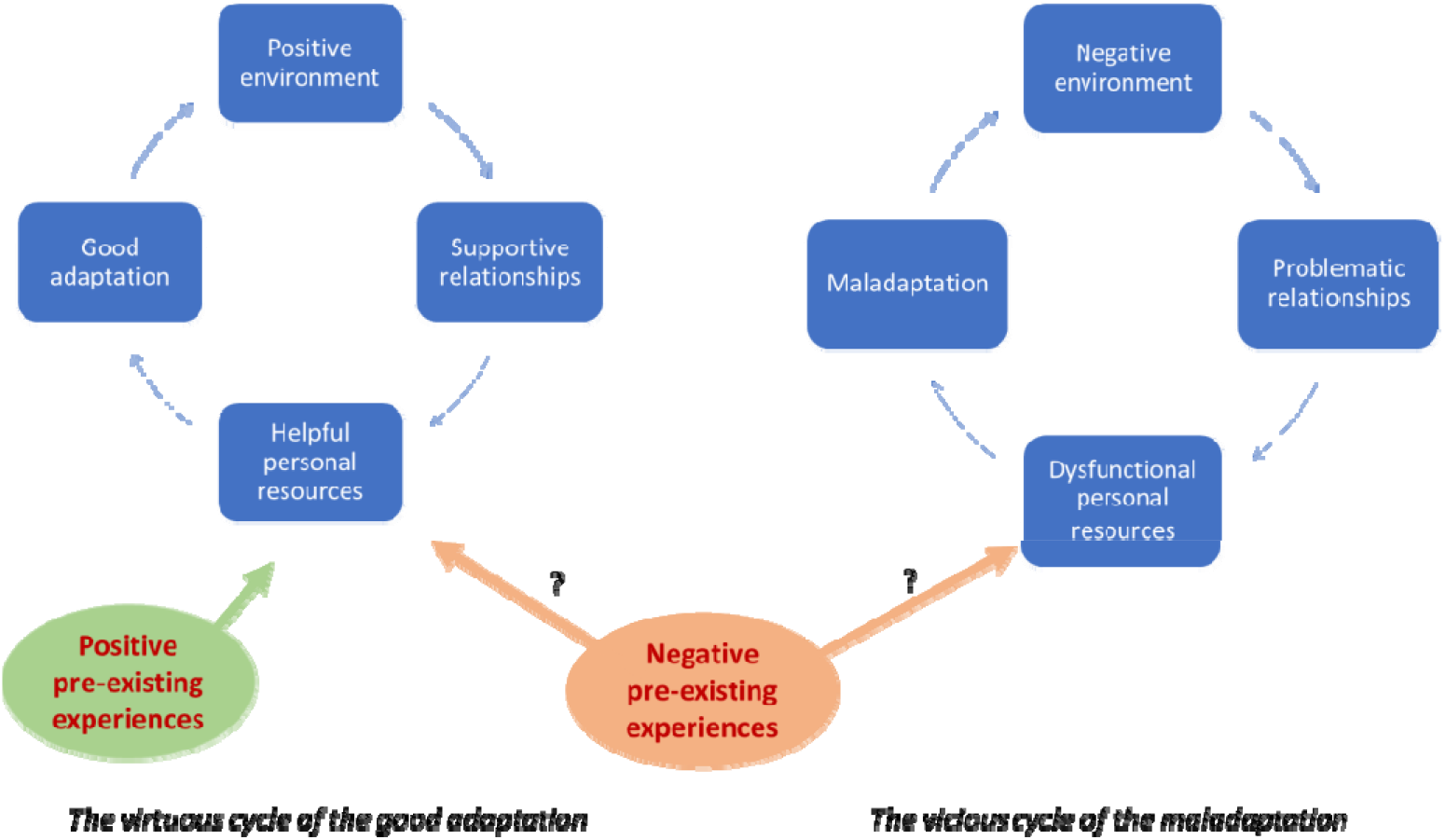
The model of Virtuous & Vicious cycles in adaptation.

Families in the virtuous cycle were able to adapt faster and more easily to the lockdown restrictions in the context of a positive family environment in which family members supported each other, had regular communication, and often spent time together doing activities. These families had their own personal resources (e.g. friendships, appropriate space at home, access to IT). They found alternative ways of doing things, reported better mental health and seemed to have adapted to ongoing challenges.

I am in a really fortunate position because obviously I have got a very supportive husband and family, you know we can all maintain contact now with technology [..], we don’t feel sorry for ourselves, we actually feel that we are really quite fortunate, and I think my girls realize that as well, how fortunate we are (family).

Families in the vicious cycle had higher levels of stress. They also experienced more difficulties following the restrictions and keeping a balance among their personal lives, childcare and work. Often, families with negative relationships had more conflicts, and lack of personal resources (e.g. lack of friendships, inadequate space at home, no access to IT). These families were less able to find alternative ways to adapt to lockdown which could lead to further stress and often denial about having to comply with the lockdown rules.

Families that don’t have kind of robust coping mechanisms, [where] parents don’t have the ability to self-regulate, struggle with how they are feeling so, you know, it is only a matter of time I think before they start to see a real kind of decline. I think as people are adjusting to the new reality that reality is going to hit them like a ton of bricks, and everything is just going to crash (social worker).

### 3.1 The role of pre-existing factors

Certain pre-existing factors and experiences had multiple impacts on peoples’ lives during lockdown, affecting their skills to adapt positively or negatively. Our analysis also revealed that families were able to recall and draw upon past events to frame their understanding of behaviour within the lockdown. Positive pre-existing factors, such as good mental health and communication within the family, helped families’ wellbeing and adherence to the lockdown measures.

We have adapted quite quickly, but I would also say this is early days and so as distancing measures have come in, and I think we are a bit lucky as well as a family we are not particularly outgoing, party loving people, so our personality and temperament has sort of protected us from the risk of these changes (family).

Negative pre-existing experiences could worsen a family’s adaptation to lockdown rules through disadvantageous conditions.

Families that I work with who have more significant complex needs or, more you know like, drug use and stuff, then they would still…that’s really impacted on the way they’ve cope (social worker).

However, negative pre-existing experiences helped some families to have resilience by using skills they developed from previous challenging experiences. These families could maintain better mental health and resilience during the whole period of lockdown and adapted better to the restrictions.

The families where at least one of the carers have had underlying health problems, they have been very quick to react, and have put in place strategies to protect themselves (social worker).

## Discussion

### Families’ experiences during COVID-19 pandemic

In line with previous studies, we found that the COVID-19 pandemic brought many changes in peoples’ life that had a strong impact on their personal and work life as well as on their health (Rizeq et al., 2021Bentenuto, 2021; Panda, 2020; Tso, 2020). The most important challenge for the families was to create and maintain a balance between parents’ work and childcare. Caregiving has been indicating as challenging during the time of major stressful events before e.g. hurricane Katrina (ref). The disruptions in families’ social contexts associated with large scale disasters may limit caregivers’ ability to create or maintain a supportive and responsive family environment and to model and guide children’s stress (Gil-Rivas, et al., 2013). As other studies found, high parental stress and the inflexibility in the environment generated by the COVID-19 pandemic increased challenges for the parental role, especially where family dysfunction and regular conflict affected both parents and children (Bentenuto, 2021; Darks, 2020 Bikmazer, 2020; Adibelli, 2020). In contrast, parental flexibility was associated with greater family cohesion, resilience, and easier adaptation to the government’s measures (Darks, 2020; Tso, 2020). … The necessity of home-schooling was a milestone challenge for most of the families, especially for parents who worked from home. Other studies found that the closing of schools and lack of social contact had a severely negative impact on children’s social and communication skills, especially for children with NDCs (Panda, 2020). Children with NDCs and special educational needs had more emotional, behavioural and social problems compared to typical development children during the lockdown (Bentenuto, 2021; Tso, 2020; Daks, 2020; Nonweiler, 2020; Bikemaker, 2020; Panda, 2020). Parents and children with NDCs, such as Autism and ADHD generally had worse wellbeing than other families during lockdown (Bentenuto, 2021; Vallejo-Slocker, 2020; Willner, 2020). This can be explained by the lack of routine, lack of immediate access to services and therapy, and limited opportunities for children to express their energy (e.g. hyperactive children), especially for those living in urban areas (Narzisi, 2020; Becker, 2020; Bentenuto, 2021; Eapen, 2020). Families with low-income, social assistance, financial problems, or history of physical and/or mental health conditions on parents, and single-parent families, faced significant struggles during lockdown (Vallejo, 2020; Tso, 2020; Daks, 2020). However, as other studies found, many parents we interviewed reported that lockdown was an opportunity to be more aware of their children’s difficulties (Panda, 2020).

Regarding the follow-up, our findings revealed that the first months of the lockdown were particularly demanding for all families (Witteveen, 2023). After the easing of the second lockdown in the UK (June 2020), we observed that time allowed some families to adapt more effectively to the restrictions. However, in some cases, the prolonged restrictions perpetuated or worsened pre-existing difficulties within families, who anticipated a return to their pre-COVID routines. At the same time, there were families who felt apprehensive about resuming their pre-COVID lives, largely due to fears of contracting the virus. While COVID-19 restrictions had a lasting psychological impact on people’s lives, research suggests that the psychological effects of not implementing quarantine and allowing disease to spread could have been even more severe (Brooks, 2020).

### The model of Virtuous & Vicious cycle

We developed the model of Virtuous and Vicious Cycles to describe the processes underlying family’s functionality and adaptation during stressful events. The Virtuous Cycle represents the consistent implementation of effective strategies to address challenges during highly stressful periods, facilitating successful adaptation. In contrast, the Vicious Cycle reflects a persistent lack of effective strategies, leading to maladaptation and increasing difficulties in coping with the guidelines.

Associations between stressors, family resources, perception of lockdown, and family adaptation have been already suggested by previous research (Bernedo et al., 2021.) Our results agree with the Double ABC-X model of family stress and coping (Reuben Hill 1983): i.e. a family’s capacity to cope with stressful situations is influenced by pile-up of stressors, family resources and perception of the situation. However, in addition, we identified the importance of “experiential learning”, i.e. a process of learning by doing (ref). The ability to learn from previous experiences was a crucial skill for families facing the challenges of the lockdown for the first time. It helped some families build resilience, enabling them to enter the virtuous cycle of adaptation while, for others, risk factors exacerbated stress, leading them into the vicious cycle of maladaptation.

Negative-preexisting experiences such as an unsupportive environment and lack of personal resources made it more difficult for families to adapt to the changes brought by the pandemic and to follow the restrictions. This is in line with the family stress model (Masarik and Conger, 2017) that elucidates how individual distress can strain family relationships and disrupt parenting, eventually threatening the health and wellbeing of children developing emotional, behavioural, and attentional difficulties (Tso, 2020; Dodd, 2020). There is evidence for a dose-response relationship between cumulative negative experiences, high stress and mental health problems in families, which can impact their coping skills during stressful events (Bøe, 2018). Thus, based on the family stress model, these difficulties could exacerbate children’s and parents’ maladjustment to the new conditions pandemic imposed (Reich et al., 2023).

In contrast, a positive pre-existing family environment, supportive relationships and personal resources helped many families, despite their high-risk status, to cope with the lockdown and adhere to government guidelines (Adibelli, 2020; Clayton, 2020). These families demonstrated higher levels of resilience that could buffer parental stress and psychosocial problems in children (Tso, 2020). Therefore, not all high-risk families with a history of stress followed the pathways as predicted by the Family Stress Model. A unique finding in our analysis was that, in some cases, negative pre-existing experiences allowed some families to develop coping mechanisms that they could later reuse in response to the stress of COVID-19. A subset of families created virtuous cycles, where they demonstrated resilience, using the opportunity of lockdown to recalibrate routines, strengthen bonds, and develop effective coping mechanisms. So, stressful events may or may not result in crises, but families could mitigate the negative effects of stressors on their well-being and functioning if they have access to appropriate resources. There may be a plausible mechanism linking (i) negative events, (ii) how families interpret them and (iii) whether or not they could transform them into “useful tools” for handling similar future events.

These contrasting findings are supported by the Adaptive Calibration Model (ACM) of stress responsivity (Del Giudice and Shirtcliff, 2011) which argues that, like other animals, we are physiologically designed to withstand stress – even extreme stressors. During the COVID-19 pandemic, families faced unprecedented challenges such as: health threats (fear of illness or death), economic stress (job losses, financial insecurity), social isolation (lack of support networks), disrupted routines (school closures, remote work). Yes these stressors influenced families to adapt in both constructive (virtuous) or destructive (vicious) ways depending on whether they were able to calibrate their coping strategies in response to these new pressures. Other authors have also found that many families during covid pandemic used their existing resources to cope with stress and to adapt to the disruptive conditions of covid-19 pandemic (Shoychet (2022). We were surprised to find that this is also true of high-risk families: one family may enter to a “vicious cycle” of adaptation, when stress amplified tensions, leading to behaviours that worsened conditions (e.g., rule-breaking led to more infections, more lockdowns, and greater stress). Another family may enter into a “virtuous cycle” of adaptation when positive adaptations reduced stress and fostered trust, creating a feedback loop of resilience (e.g., following guidelines reduced infections, eased lockdowns, and building community trust).

### Suggestions for future public health crises

Often public stressful situations such as war, natural disaster, pandemic etc. disrupt the multiple layers of the social fabric that support families’ wellbeing (Gil-Rivas, et al., 2013). However, during a public crisis, supporting the interaction between families’ ecosystems (Bronfenbrenner, 1977) and sharing personal experiences, could potentially increase resilience by providing new or enhanced personal and social resources for families, (Slone and Peers, 2021; Grey et al., 2018). In the case of Covid-19 pandemic, it was an unfamiliar stressful event that could be viewed as multifactorial reflecting the interaction of the micro-system of a nuclear family’s and macro systems of national policies. Worldwide, government and public health organisations trying to mitigate the spread of COVID-19 implemented various public health measures such as lockdowns and closures of businesses and schools, traveling restrictions, mandatory masks etc., though, the effectiveness of these guidelines was affected by the heterogeneity of the needs in the population (Talic, et al., 2021). Our study showed the importance, in future societal crises, of helping families understand the ways in which they have coped with stress in the past and encouraging families to use these pre-existing coping strategies to meet the new challenges they are facing. Specifically, based on our findings and previous research, we support that public policies established by the government to deal with public stressful events such as pandemic, war or natural disasters, should be developed by focusing on increasing existing skills and knowledge in families rather than trying to teach new ones. High-risk families, those with history of disadvantages, physical and/or mental health issues, children with learning disabilities and NDCS or foster-care, may struggle to process new information during a crisis. Building on existing skills should be a central focus to create individual and societal resilience.

## Implications and Future Directions

These findings emphasize the importance of recognizing both vulnerability and resilience within high-risk families and challenging the assumption that all families under stress are inevitably at risk of a downward spiral. Instead, by understanding the conditions under which some families transition to a virtuous cycle, interventions can be better tailored to support adaptive behaviours that promote resilience. Future research should continue to explore these dynamics longitudinally, identifying specific factors that differentiate families who enter virtuous cycles from those whose functioning worsens. This understanding will be crucial for developing targeted support systems and policies that not only address immediate needs but also foster long-term resilience.

## Limitations

Although this study is important from both clinical and research perspectives, some limitations should be considered. We made efforts to include a wide range of high-risk families, especially those families unlikely to participate in surveys, however, we may still have excluded people without IT equipment and limited internet access. We recruited families and professionals only from Scotland and England so, our findings may not be generalisable to other populations.

## Conclusion

Our findings present the challenges UK families faced during a major societal stressor. We introduce the model of virtuous and vicious cycles in family coping and the contribution – both positive and negative -of pre-existing stressful experiences. By recognizing and supporting these adaptive processes, researchers and practitioners can better support family systems in the face of significant stressors, promoting a pathway to resilience rather than vulnerability.

## Data Availability

All data produced in the present study are available upon reasonable request to the authors

